# Urban Heat Islands and their Effect on Incident Myocardial Infarctions

**DOI:** 10.1101/2025.04.04.25325284

**Authors:** Brenna Lilley, Olawale Amubieya, Boback Ziaeian

**Author notes:** Corresponding Author: Brenna Lilley, MD Attending Physician, Department of Hospital Medicine, Kaiser Permanente West Los Angeles Medical Center Mailing Address: 6041 Cadillac Ave, Los Angeles, CA 90034.

## Abstract

**Background:** Climate change is causing an increasing number of extreme heat events, which are associated with a higher risk of cardiovascular disease (CVD)-related morbidity and mortality. Urban areas are particularly susceptible to extreme heat due to the urban heat island (UHI) effect.

**Objectives:** This study investigates the effect of UHI exposure on clinically significant CVD and how this relationship changes when accounting for measures of social vulnerability.

**Methods:** We used publicly available data from California urban areas on a census-tract level, including the UHI index (UHII), CVD incidence as measured by the age-adjusted rate of acute myocardial infarction (MI), the Social Vulnerability Index (SVI), and racial and ethnic group percentages. We performed regression analyses, including simple linear regression, partial regression, and multiple regression with interaction terms, to model these relationships. We created census-tract level bivariate choropleth maps of urban areas in California counties.

**Results:** The regression models estimate that residents in census tracts with a mean UHII have a 22% increased risk of acute MI and residents in census tracts with the highest UHII have a 112% increased risk of acute MI when compared to residents living in census tracts with the lowest UHII. Additionally, for any given value of UHII, the rate of acute MI was predicted to be higher in census tracts with higher percent people of color.

**Discussion:** This study highlights the intersection of UHII, CVD incidence, and social vulnerability within urban areas on a census-tract level. It emphasizes the need to implement mitigation strategies to decrease the UHI effect, thereby preventing heat-related CVD events.

## Introduction

Climate change has not only led to a gradual increase in global average temperature, but also increasing frequency, duration, and intensity of extreme heat events including heat waves.^1,2^ These extreme heat events have been associated with increased cardiovascular disease (CVD)-related morbidity and mortality.^3–6^

Urban areas are particularly vulnerable, as they can develop higher temperatures compared to their rural counterparts, a phenomenon known as the urban heat island (UHI) effect. UHIs are caused by increased heat-absorptive surfaces and heat-generating activities in combination with decreased evaporative cooling from vegetation.^7^ Extreme heat events can intensify UHIs, causing increased heat stress in urban environments.^8^ During extreme heat events, urban areas with higher UHI intensity have been shown to have increased CVD-related morbidity and mortality.^6,9^

Other factors that contribute to increased risk for CVD include race and ethnicity. Individuals who identify as Hispanic, non-Hispanic Black, and American Indian have higher rates of CVD or predisposing conditions such as hypertension, obesity, and diabetes.^10–11^ People of color (POC, individuals who do not identify as non-Hispanic white) experience health disparities due to social determinants of health, which include unhealthy living conditions caused by the built environment.^11^ Given the health risks associated with heat stress, living in an area with higher UHI exposure can be considered as an unhealthy living condition. Data has shown that both POC, regardless of socioeconomic status, and people living below the poverty line live in areas with increased UHI exposure.^12^

As described above, CVD risk is impacted by various social and structural factors, including race, ethnicity, and heat exposure. The relationship between extreme heat exposure, CVD, and race and ethnicity has been investigated on county-level analyses, showing greater mortality rates in non-Hispanic Blacks.^4^ Since there is significant intra-city variation in UHI intensity, it is important to analyze data on a geographic scale that is higher-resolution than the county, such as the census-tract level, to capture such intricacies.^12^ Additionally, studies have not yet investigated how baseline UHI exposure, regardless of extreme heat events, impacts clinically significant CVD. This study aims to investigate the relationships between UHI intensity, CVD, and social vulnerability on a census-tract level within urban areas in the state of California.

## Methods

We received IRB exemption from UCLA for this study.

### Data Sources

To evaluate the relationship between UHI intensity and CVD incidence, we used publicly available data from CalEPA and CalEnviroScreen 4.0, respectively. We also used data from the CDC/ATSDR for measures of social vulnerability and race and ethnicity.

#### Urban Heat Island Index

The primary exposure of interest was the UHI index (UHII), calculated by CalEPA. UHII is defined as the “positive temperature differential over time between an urban census tract and nearby upwind rural reference points at a height of two meters above ground level, where people experience heat,” (CalEPA, 2023). The UHII is derived from measurements over 182 warm season days from 2006 to 2013 in California urban areas for each census tract.^8^ The data for the measurements were obtained from the National Center for Environmental Prediction, National Center for Atmospheric Research, National Weather Service, and National Oceanic and Atmospheric Administration.^8^ UHII is reported as degree-hours per day on a Celsius scale, creating an index that combines both the intensity and duration of heat attributed to the UHI effect.^7^

#### Cardiovascular Disease Incidence

The primary outcome of interest was CVD incidence obtained from the CalEnviroScreen 4.0 dataset. CVD incidence was derived from a spatially modeled, age-adjusted rate of emergency department visits in California for acute myocardial infarction (MI) per 10,000 residents per year averaged over 2015 to 2017. The acute MI data was obtained from Emergency Department and Patient Discharge Datasets from the State of California, Office of Statewide Health Planning and Development. The CVD incidence is a census tract estimate, derived from population-weighted 2010 census block estimates.^13^

#### Social Vulnerability Index

A secondary exposure of interest that also served as a control variable was the Social Vulnerability Index (SVI) created by CDC/ATSDR. The SVI is a place-based index designed to identify communities experiencing social vulnerability.^14^ The 2018 SVI database was used in this study as it was derived from 2010 census tracts and demographic variables from the 2014-2018 American Community Survey (ACS).^14^ The California data for overall SVI and the SVI Themes (Theme 1: Socioeconomic Status, Theme 2: Household Composition and Disability, Theme 3: Minority Status and Language, and Theme 4: Housing Type and Transportation) were used in the analysis and range from 0.0 to 1.0. See Table S1 for the demographic variables from ACS that were included in the 2018 SVI database.

#### Race and Ethnicity

Another secondary exposure of interest was race and ethnicity, represented in this study as percent POC (% POC). This was derived from the 2018 SVI database variable “percentage minority estimate”, which was defined as all persons except white, non-Hispanic based on data from ACS 2014-2018.^14^

### Statistical Analysis

The datasets were organized by census-tract FIPS codes. Data for CVD incidence, SVI, and % POC with FIPS codes that corresponded to urban areas with a calculated UHII were included.

#### Primary Analysis: CVD Incidence as a Function of UHII

CVD incidence and UHII were plotted in a bivariate distribution (Figure 1). The data was modeled with a natural cubic spline with four knots using default quantiles. Areas of the spline between the inflection points were divided to perform regression analysis. To find the values of UHII at these inflection points, the spline was evaluated over a fine grid to approximate the derivative using finite differences and the values of UHII which produced the smallest absolute derivate of the spline were calculated. These values were UHII of 22 and 79 DH/day, representing the values of UHII at the inflection points. Simple linear regression was performed for CVD incidence as a function of UHII for all three UHII groups: UHII < 22 DH/day (UHII Group 1), UHII 22-78.9 DH/day (UHII Group 2), and UHII ≥ 79 DH/day (UHII Group 3).

**Figure 1.**
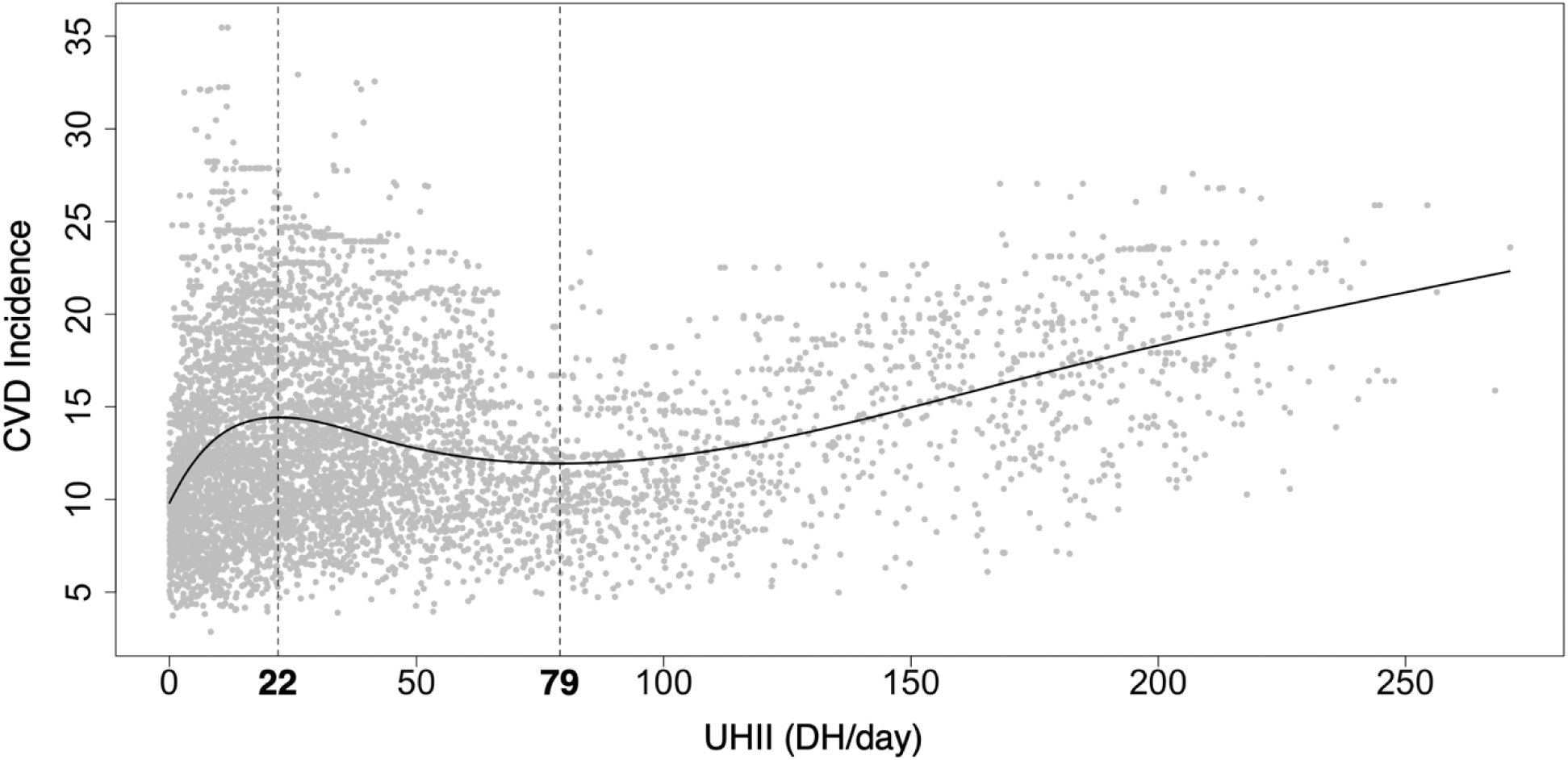
CVD incidence as a function of UHII. CVD incidence is the age-adjusted rate of emergency department visits in California for acute MI per 10,000 residents per year averaged over 2015-2017. Modeling was performed with a natural cubic spline. The values of UHII at the inflection points were determined to be UHII 22 and 79 DH/day. To perform regression analysis, the data was split into UHII groups using the inflection points as cutoffs: UHII < 22 DH/day (UHII Group 1), UHII 22-78.9 DH/day (UHII Group 2), and UHII ≥ 79 DH/day (UHII Group 3).

#### Secondary Analyses

We used simple linear regression to analyze the bivariate relationships of CVD incidence and SVI, UHII and SVI, CVD incidence and % POC, and UHII and % POC by census-tract. To determine how SVI impacts the relationship between CVD incidence and UHII, we performed partial regression analysis for CVD incidence as a function of UHII while controlling for SVI. We used multiple regression with an interaction term to model the effect of the interaction between UHII and % POC on CVD incidence. We included the minimum, first through third quartiles, and maximum values of % POC for each UHII group in the multiple regression models. For the regression analyses, all *p* values were two-sided, and we considered all values of *p* < 0.05 as statistically significant. For linear regression analysis and partial regression analysis, we calculated Pearson correlation coefficients (*r*) and qualified the strength of the relationship by the Chan YH method.^15^ For multiple regression analysis with an interaction term, we calculated adjusted *R*^2^ values. All statistical analyses were performed in RStudio. Natural cubic spline was done using the splines package. Partial regression analysis was done with the patchwork package. Multiple regression analysis with an interaction term was done using the sjPlot package.

### Choropleth Maps

For geographical visualization of the data, we created bivariate choropleth maps in RStudio using the biscale package. 2010 census tract Shapefiles were downloaded from census.gov and converted to geoJSON files, which were imported as map boundaries into RStudio. For the Los Angeles County choropleth map, the county boundary was downloaded from Los Angeles GeoHub and the basemap was created on maps.stamen.com using the terrain background.

## Results

The analysis included 6,323 census tracts within urban areas across the state of California. There were 2,573 census tracts within the UHII < 22 DH/day group (UHII Group 1), 2,574 census tracts within the UHII 22-78.9 DH/day group (UHII Group 2), and 1,176 census tracts within the UHII ≥ 79 DH/day group (UHII Group 3).

Table 1 shows the census tract characteristics across all values of UHII and UHII Groups 1-3. UHII ranged from 0.03 – 271.16 DH/day with a mean of 46.89 DH/day. For all values of UHII, the mean CVD incidence rate was 13.43 per 10,000 residents per year. Mean CVD incidence and mean % POC were lowest in UHII Group 1 and highest in UHII Group 3.

**Table 1:** Characteristics of census tracts within urban areas across California.

|  | All UHII | UHII Group 1 | UHII Group 2 | UHII Group 3 |
| --- | --- | --- | --- | --- |
| **Census tracts, #** | 6,323 | 2,573 | 2,574 | 1,176 |
| **UHII, DH/day** |  |  |  |  |
| Range | 0.03 – 271.16 | 0.03 – 21.99 | 22.00 – 78.90 | 79.00 – 271.16 |
| Mean (IQR) | 46.89 (47.19) | 9.18 (15.54) | 41.99 (22.92) | 140.08 (78.31) |
| **CVD Incidence** |  |  |  |  |
| Mean (IQR) | 13.43 (7.21) | 12.87 (7.03) | 13.53 (6.64) | 14.44 (7.34) |
| **Overall SVI** |  |  |  |  |
| Mean (IQR) | 0.51 (0.51) | 0.48 (0.54) | 0.53 (0.52) | 0.52 (0.44) |
| **POC, %** |  |  |  |  |
| Mean (IQR) | 64.5 (44.0) | 61.5 (45.5) | 65.8 (45.3) | 68.2 (36.7) |

UHII, created by CalEPA, was averaged over 182 warm season days from 2006-2013 and represented as degree hours per day (DH/day). UHII was divided into three groups based on inflection points in the bivariate distribution of UHII and CVD incidence. CVD incidence, from CalEnviroScreen 4.0, is the spatially modeled, age-adjusted rate of emergency department visits in California for acute myocardial infarction (MI) per 10,000 residents per year averaged over 2015-2017. Overall SVI and % POC were obtained from the 2018 SVI database and derived from the 2014-2018 American Community Survey.

### Primary Analysis

#### CVD Incidence and UHII

Simple linear regression analysis and models of CVD incidence rate and UHII Groups 1-3 are shown in Table 2 and Figure 2. For UHII Group 1, there was a significant positive relationship between UHII and CVD incidence rate, with a 0.197 event (95% CI: 0.166, 0.228) increase in CVD incidence per 10,000 per year for every 1 DH/day increase in UHII and a poor positive correlation (Table 2, Figure 2A). For UHII Group 2, there was a significant negative relationship between UHII and CVD incidence rate, with a 0.045 event (95% CI: -0.057, -0.033) decrease in CVD incidence per 10,000 per year for every 1 DH/day increase in UHII and a poor negative correlation (Table 2, Figure 2B). For UHII Group 3, there was a significant positive relationship between UHII and CVD incidence rate, with a 0.066 event (95% CI: 0.060, 0.072) increase in CVD incidence per 10,000 per year for every 1 DH/day increase in UHII and a moderate positive correlation (Table 2, Figure 2C).

**Table 2:**
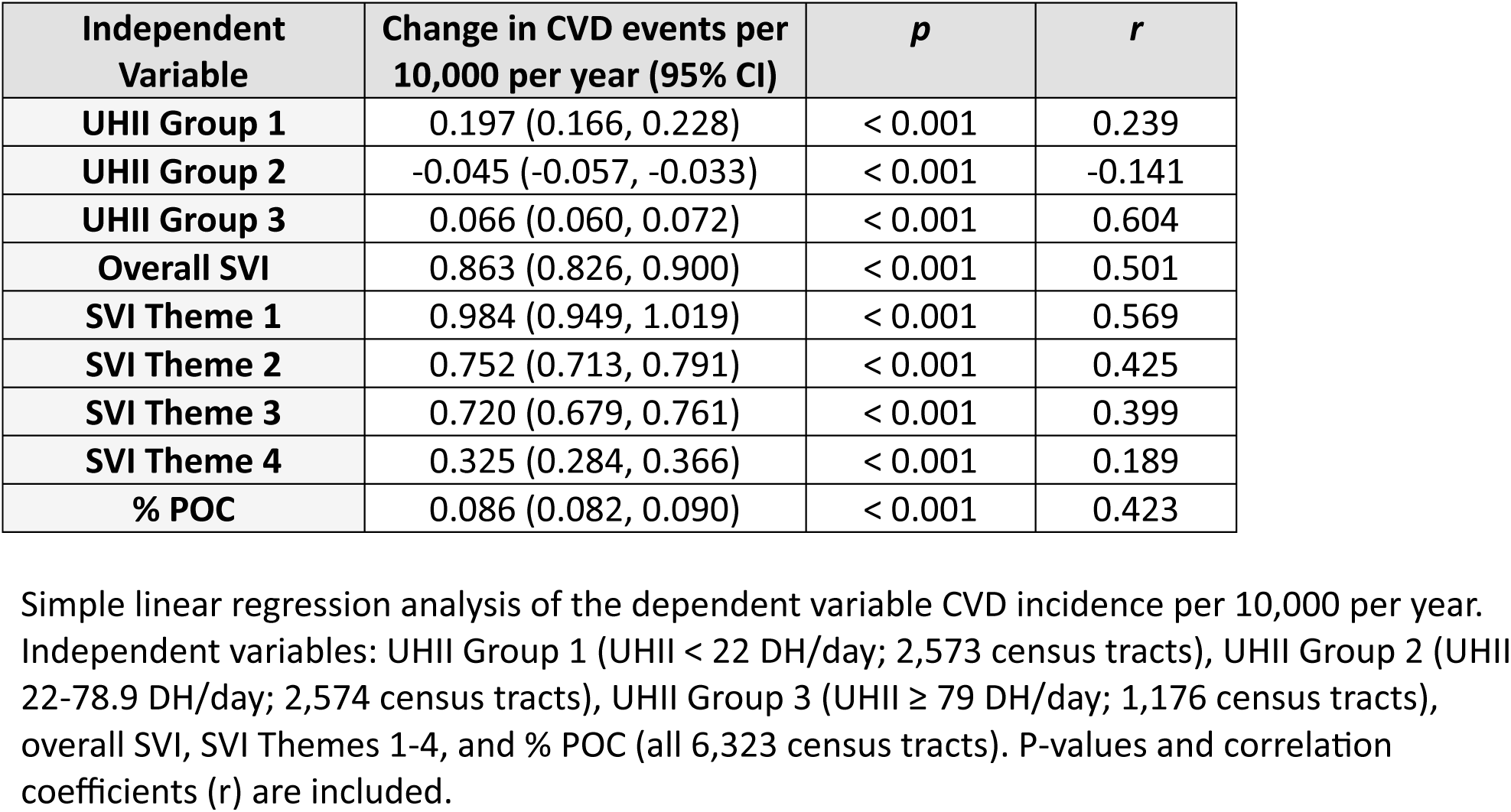
Change in CVD incidence rate related to UHII, SVI, and POC on a census-tract level within urban areas in California.

| Independent Variable | Change in CVD events per 10,000 per year (95% CI) | *p* | *r* |
| --- | --- | --- | --- |
| **UHII Group 1** | 0.197 (0.166, 0.228) | < 0.001 | 0.239 |
| **UHII Group 2** | -0.045 (-0.057, -0.033) | < 0.001 | -0.141 |
| **UHII Group 3** | 0.066 (0.060, 0.072) | < 0.001 | 0.604 |
| **Overall SVI** | 0.863 (0.826, 0.900) | < 0.001 | 0.501 |
| **SVI Theme 1** | 0.984 (0.949, 1.019) | < 0.001 | 0.569 |
| **SVI Theme 2** | 0.752 (0.713, 0.791) | < 0.001 | 0.425 |
| **SVI Theme 3** | 0.720 (0.679, 0.761) | < 0.001 | 0.399 |
| **SVI Theme 4** | 0.325 (0.284, 0.366) | < 0.001 | 0.189 |
| **% POC** | 0.086 (0.082, 0.090) | < 0.001 | 0.423 |

Simple linear regression analysis of the dependent variable CVD incidence per 10,000 per year. Independent variables: UHII Group 1 (UHII < 22 DH/day; 2,573 census tracts), UHII Group 2 (UHII 22-78.9 DH/day; 2,574 census tracts), UHII Group 3 (UHII ≥ 79 DH/day; 1,176 census tracts),
overall SVI, SVI Themes 1-4, and % POC (all 6,323 census tracts). P-values and correlation coefficients (r) are included.

**Figure 2.**
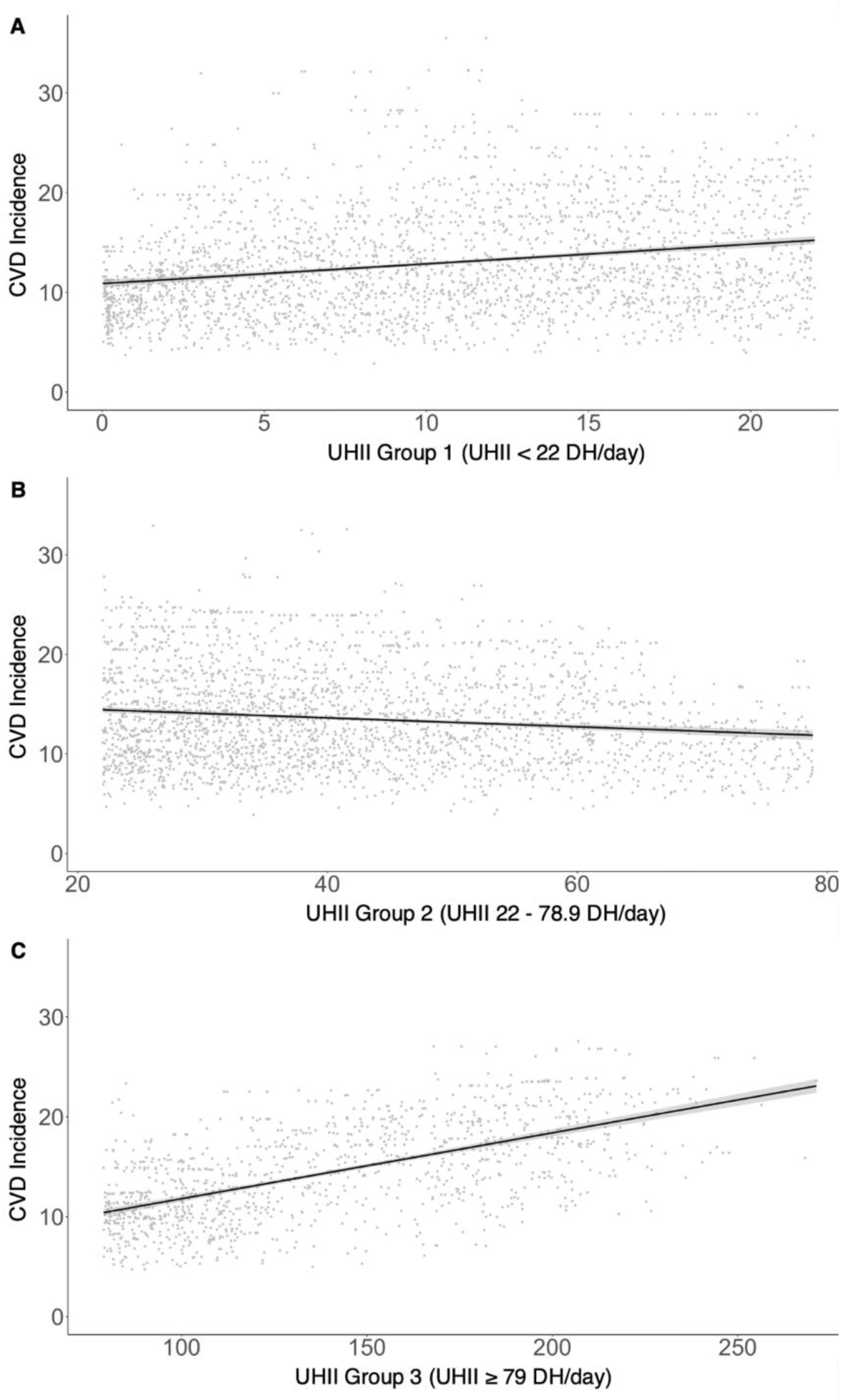
Simple linear regression analysis of CVD incidence and UHII. Linear regression models for UHII Groups 1 (A), 2 (B), and 3 (C) superimposed on scatterplots of CVD incidence per 10,000 per year and UHII; see Table 2 for simple linear regression coefficients, P values, and r values.

The simple linear regression models estimate that for a census tract with the lowest UHII (0.03 DH/day), the CVD incidence rate will be 10.89 per 10,000 per year. For a census tract with the mean UHII (46.89 DH/day), the CVD incidence rate will be 13.31 per 10,000 per year with a relative risk ratio of 1.22 when comparing to a census tract with the lowest UHII. For a census tract with the highest UHII (271.16 DH/day), the CVD incidence rate will be 23.08 per 10,000 per year with a relative risk ratio of 2.12 when comparing to a census tract with the lowest UHII.

### Secondary Analyses

#### CVD Incidence and SVI

Simple linear regression analyses of CVD incidence rate, overall SVI, and the SVI Themes are shown in Table 2. Simple linear regression model of CVD incidence rate and overall SVI is shown in Figure 3A and models of CVD incidence rate and SVI Themes are shown in Figure S1. There was a significant positive relationship between CVD incidence rate and overall SVI with a 0.863 event (95% CI: 0.826, 0.900) increase in CVD incidence per 10,000 per year for every 0.1 unit increase in overall SVI and a fair positive correlation (Table 2, Figure 3A). Further analyses were done using SVI Themes 1-4. There was a significant positive relationship between CVD incidence rate and SVI Theme 1, with a 0.984 event (95% CI: 0.949, 1.019) increase in CVD incidence per 10,000 per year for every 0.1 unit increase in SVI Theme 1 and a moderate positive correlation (Table 2, Figure S1A). There was a significant positive relationship between CVD incidence rate and SVI Theme 2, with a 0.752 event (95% CI: 0.713, 0.791) increase in CVD incidence per 10,000 per year for every 0.1 unit increase in SVI Theme 2 and a fair positive correlation (Table 2, Figure S1B). There was a significant positive relationship between CVD incidence rate and SVI Theme 3, with a 0.720 event (95% CI: 0.679, 0.761) increase in CVD incidence per 10,000 per year for every 0.1 unit increase in SVI Theme 3 and a fair positive correlation (Table 2, Figure S1C). There was a significant positive relationship between CVD incidence rate and SVI Theme 4, with a 0.325 event (95% CI: 0.284, 0.366) increase in CVD incidence per 10,000 per year for every 0.1 unit increase in SVI Theme 4 and a poor positive correlation (Table 2, Figure S1D).

**Figure 3.**
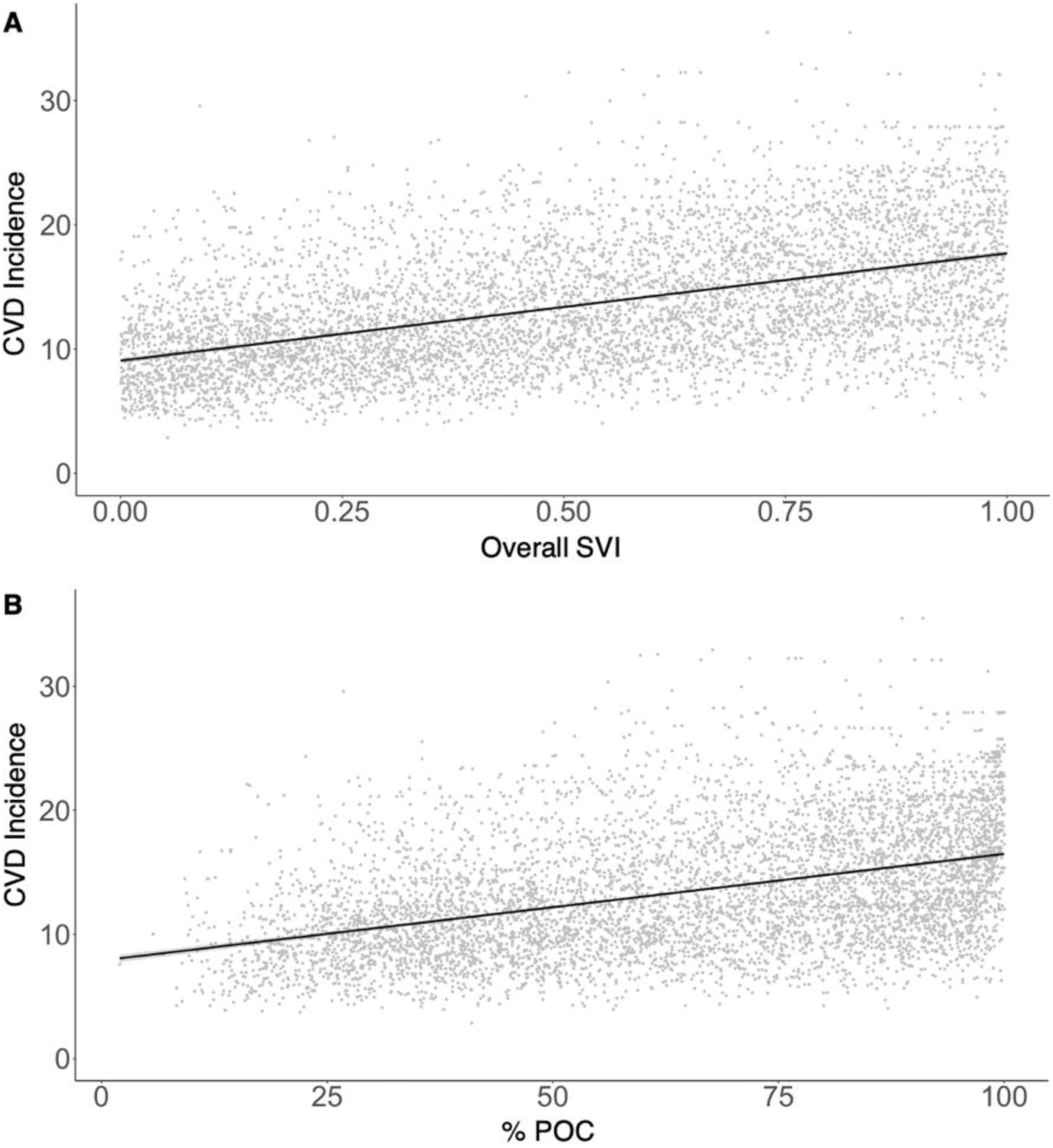
Simple linear regression analysis of CVD incidence, SVI, and % POC. Linear regression model of CVD incidence per 10,000 per year and overall SVI (A); linear regression model of CVD incidence and % POC (B); see Table 2 for simple linear regression coefficients, P values, and r values.

#### UHII and SVI

There was no significant linear relationship between UHII and overall SVI or SVI Themes 1-4 in a bivariate distribution.

#### CVD Incidence and UHII, Controlling for SVI

Partial regression analysis and models of CVD incidence rate as a function of UHII when controlling for overall SVI are shown in Table 3 and Figure 4. When controlling for overall SVI, there was still a significant positive relationship between CVD incidence rate and UHII Group 1, with a 0.053 event (95% CI: 0.026, 0.080) increase in CVD incidence per 10,000 per year for every 1 DH/day increase in UHII and a poor positive correlation (Table 3, Figure 4A). There was still a significant negative relationship between CVD incidence rate and UHII Group 2, with a 0.022 event (95% CI: -0.034, -0.010) decrease in CVD incidence per 10,000 per year for every 1 DH/day increase in UHII and a poor negative correlation (Table 3, Figure 4B). There was still a significant positive relationship between CVD incidence rate and UHII Group 3, with a 0.060 event (95% CI: 0.056, 0.064) increase in CVD incidence per 10,000 per year for every 1 DH/day increase in UHII and a moderate positive correlation (Table 3, Figure 4C).

**Table 3.** Change in CVD incidence rate per 1 DH/day change in UHII when controlling for SVI on a census-tract level in California urban areas.

| Independent Variable | Control | Change in CVD events per 10,000 per year (95% CI) | *p* | *r* |
| --- | --- | --- | --- | --- |
| UHII Group 1 | Overall SVI | 0.053 (0.026, 0.080) | < 0.001 | 0.076 |
|  | SVI Theme 1 | 0.028 (0.003, 0.053) | 0.031 | 0.042 |
|  | SVI Theme 2 | 0.092 (0.065, 0.119) | < 0.001 | 0.125 |
|  | SVI Theme 3 | 0.090 (0.061, 0.119) | < 0.001 | 0.121 |
|  | SVI Theme 4 | 0.177 (0.146, 0.208) | < 0.001 | 0.218 |
| UHII Group 2 | Overall SVI | -0.022 (-0.034, -0.010) | < 0.001 | -0.075 |
|  | SVI Theme 1 | -0.017 (-0.029, -0.005) | 0.002 | -0.060 |
|  | SVI Theme 2 | -0.035 (-0.047, -0.023) | < 0.001 | -0.118 |
|  | SVI Theme 3 | -0.031 (-0.043, -0.019) | < 0.001 | -0.102 |
|  | SVI Theme 4 | -0.037 (-0.049, -0.025) | < 0.001 | -0.119 |
| UHII Group 3 | Overall SVI | 0.060 (0.056, 0.064) | < 0.001 | 0.582 |
|  | SVI Theme 1 | 0.053 (0.049, 0.057) | < 0.001 | 0.539 |
|  | SVI Theme 2 | 0.060 (0.054, 0.066) | < 0.001 | 0.552 |
|  | SVI Theme 3 | 0.068 (0.064, 0.072) | < 0.001 | 0.630 |
|  | SVI Theme 4 | 0.065 (0.059, 0.071) | < 0.001 | 0.604 |

Partial regression analysis of the dependent variable CVD incidence per 10,000 per year as a function of the independent variable UHII when controlling for overall SVI and SVI Themes 1-4. UHII was divided into groups: UHII Group 1 (UHII < 22 DH/day; 2,573 census tracts), UHII Group 2 (UHII 22-78.9 DH/day; 2,574 census tracts), and UHII Group 3 (UHII ≥ 79 DH/day; 1,176 census tracts). P-values and correlation coefficients (r) are included.

**Figure 4.**
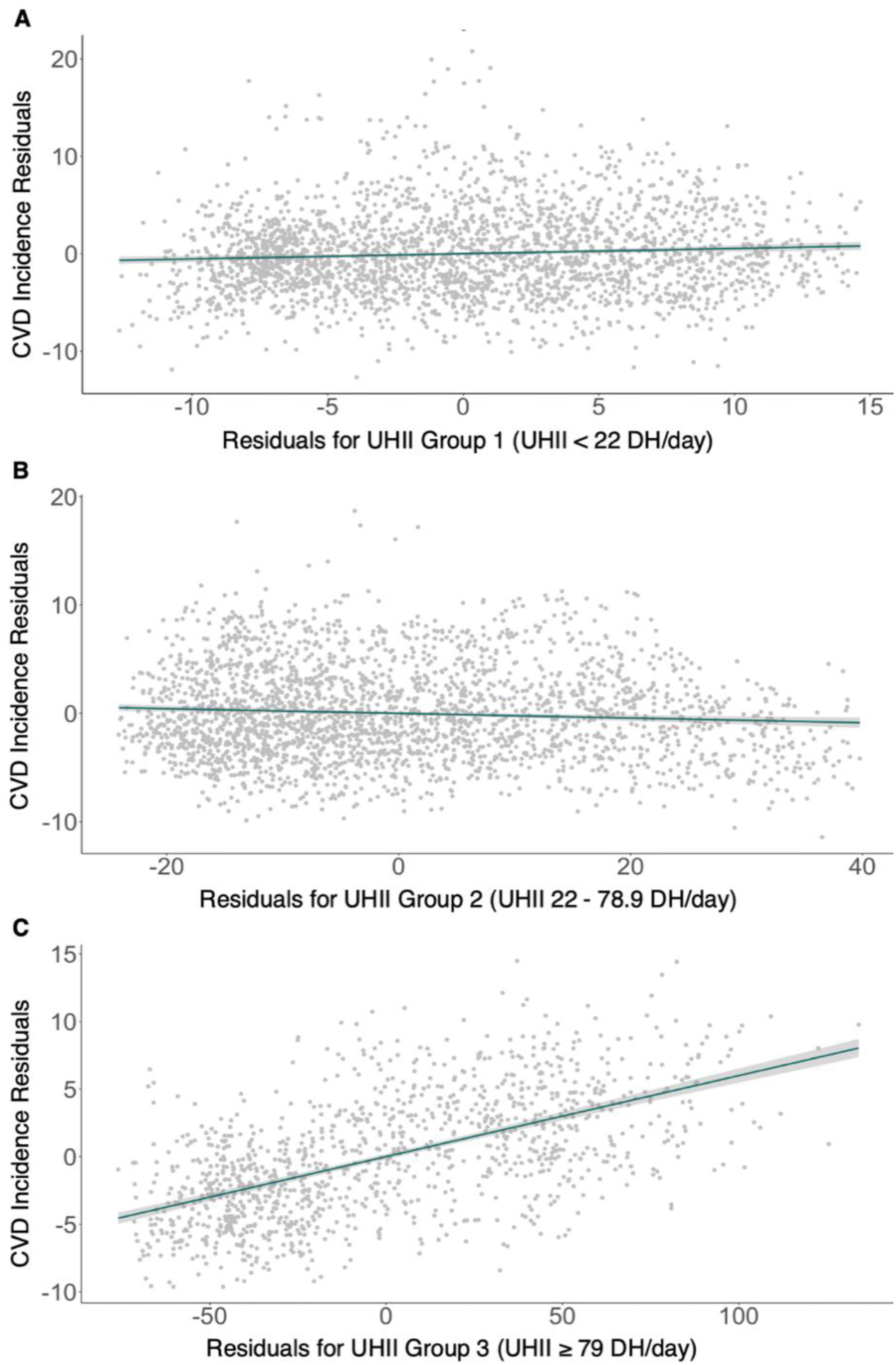
Partial regression analysis of CVD incidence as a function of UHII when controlling for overall SVI. Partial regression models using UHII Groups 1 (A), 2 (B), and 3 (C); Y-axis = residuals from regressing CVD incidence per 10,000 per year against overall SVI; X-axis = residuals from regressing UHII against overall SVI; see Table 3 for partial regression coefficients, P values, and r values.

Partial regression analysis and models of CVD incidence rate as a function of UHII when controlling for SVI Themes 1-4 are shown in Table 3 and Figures S2-S4. The linear relationships remained significant and did not change directionality.

#### CVD Incidence and % POC

Simple linear regression analysis and model of CVD incidence rate and % POC are shown in Table 2 and Figure 3B. There was a significant positive relationship between CVD incidence rate and % POC, with a 0.086 event (95% CI: 0.082, 0.090) increase in CVD incidence per 10,000 per year for every 1% increase in POC. There was a fair positive correlation between CVD incidence and % POC.

#### UHII and % POC

There was no significant linear relationship between UHII and % POC in a bivariate distribution.

#### CVD Incidence, UHII, and % POC

Multiple regression models with interaction terms are shown in Figure 5. For each UHII group, five models are portrayed using different values of % POC (the minimum, first through third quartiles, and maximum values of % POC within the UHII group). When using different values of % POC in the regression model, the relationship between UHII and CVD incidence is altered, as evident by changes in slopes and intercepts. For all UHII groups, there was a statistically significant interaction effect when modeling CVD incidence rate as a function of the interaction between UHII and % POC (Table 4).

**Figure 5.**
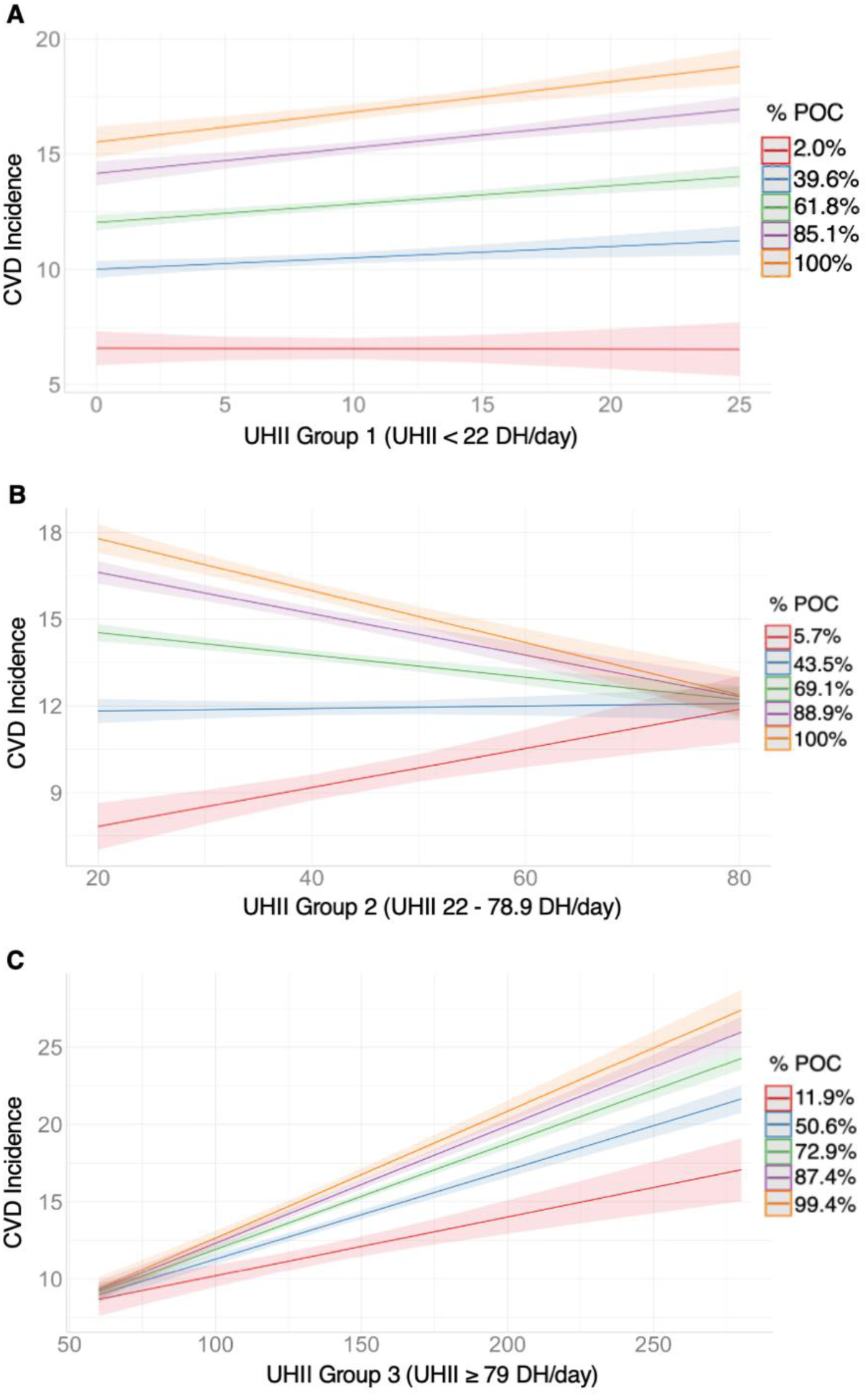
Multiple regression analysis of CVD incidence and the interaction between UHII and % POC. Multiple regression models with interaction terms shown for UHII Groups 1 (A), 2 (B), and 3 (C); values of % POC are the minimum (red), first quartile (blue), second quartile (green), third quartile (purple), and maximum (orange) values of % POC for each UHII group; see Table 4 for interaction coefficients, P values, and R^2^ values.

**Table 4.**
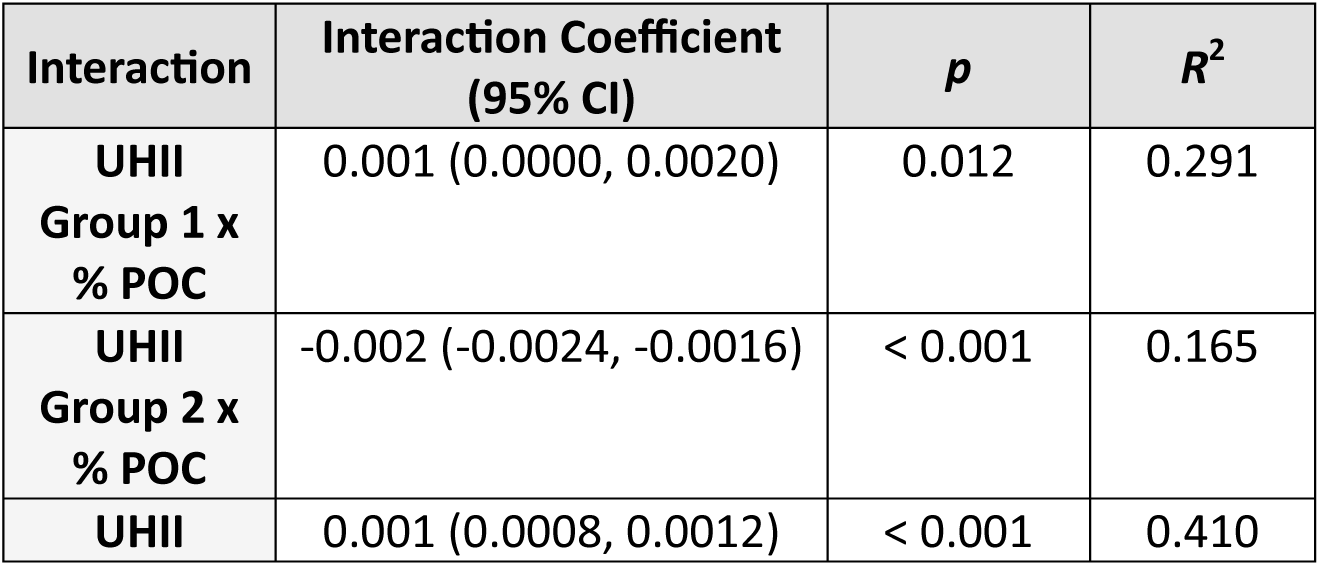
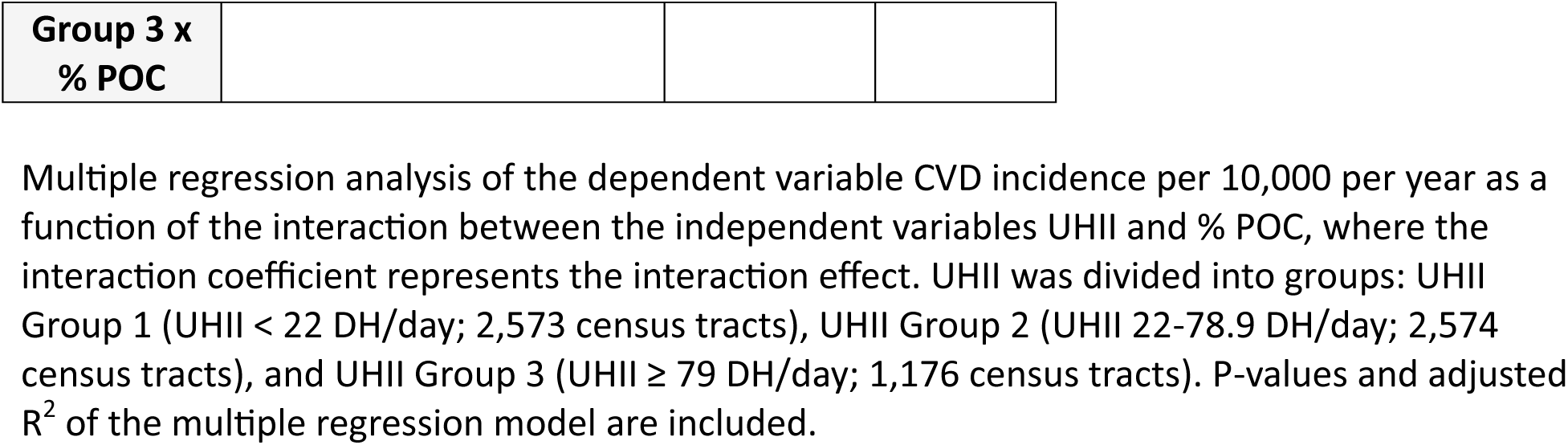
Interaction effect between UHII and % POC on a census-tract level in California urban areas.

### Choropleth Maps

Bivariate choropleth maps were created for urban areas of several of the most populous California counties, illustrating the relationships between CVD incidence rate and UHII, % POC and UHII, and CVD incidence rate and overall SVI. Figure 6 demonstrates the choropleth maps of Los Angeles County. Other county maps are presented in Figures S5-S11.

**Figure 6.**
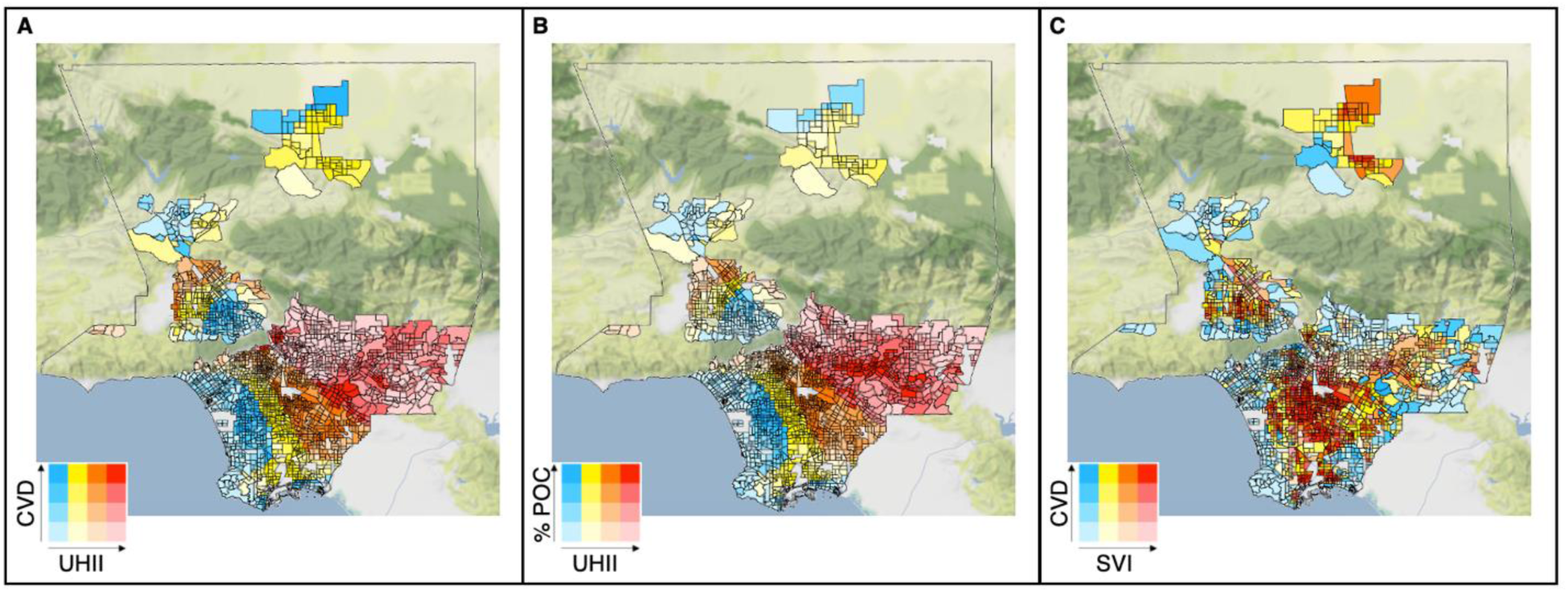
Bivariate choropleth maps of urban areas in Los Angeles County. CVD incidence per 10,000 per year and UHII (A), % POC and UHII (B), and CVD incidence and overall SVI (C). For other counties in California, see Figure S5-11.

## Discussion

The mean CVD incidence, as measured by the age-adjusted rate of acute MI per 10,000 residents per year, was lowest in the UHII < 22 DH/day group and highest in the UHII ≥ 79 DH/day group across urban areas in California. The regression models estimate that for a given census tract with the mean UHII of 46.89 DH/day, there will be a rate of 13.31 acute MI per 10,000 residents per year; these residents have a 22% increased risk of acute MI compared to residents living in a census tract with the lowest UHII (0.03 DH/day). For a given census tract with the highest UHII of 271.16 DH/day, the rate increases to 23.08 acute MI per 10,000 residents per year and residents will have a 112% increased risk of acute MI.

When investigating the relationship between social vulnerability and CVD, increasing SVI was associated with increasing CVD incidence. This is consistent with the framework that social determinants of health contribute to the development of CVD.^11^ The strongest correlation was seen between CVD incidence and socioeconomic status (SVI Theme 1). Even when controlling for social vulnerability, the relationships between CVD incidence and UHII remained significant.

On a census tract level, increasing % POC in the population was associated with increasing CVD incidence. This is consistent with evidence that certain racial and ethnic groups are more likely to develop CVD.^10,11^ Mean % POC was lowest in the UHII < 22 DH/day group and highest in the UHII ≥ 79 DH/day group, supporting prior work showing that POC are more likely to live in areas with increased UHI exposure.^12^ Multiple regression analyses showed that the effect of UHII on CVD incidence is dependent on % POC and that for any given value of UHII, the CVD incidence is predicted to be higher in census tracts with higher % POC.

Several studies have investigated the effect of extreme heat events on CVD. A systemic review and meta-analysis found a positive relationship between increasing temperature and CVD-related morbidity and mortality.^3^ A U.S. county-level longitudinal analysis showed that each additional day of extreme heat was associated with a 0.12% increased monthly CVD-related mortality rate.^4^ A systematic review and meta-analysis revealed that exposure to heat waves was associated with an increased risk of MI.^5^ A time series analysis across U.S. urban areas demonstrated that extreme heat increased the risk of CVD-related hospitalization by 1.5%.^6^ Additionally, other studies have illustrated the impact of UHIs on CVD during periods of extreme heat: in U.S. urban areas, the risk of CVD hospitalization due to extreme heat was greater in areas with higher UHI intensity, and in Beijing, hotter temperatures caused by the UHI effect were associated with increased CVD mortality. ^6,9^

This study further supports the association between heat exposure and increased risk of CVD, specifically acute MI. It also offers a novel perspective by focusing on the effect of the baseline relative increase in temperature attributed to UHIs and does not fixate on periods of extreme heat. This study suggests that census tracts with higher UHI intensity are associated with increased CVD incidence regardless of extreme heat events.

One of the strengths of this study was using census-tract level data, underscoring the range of UHII, CVD incidence, race and ethnicity, and social vulnerability across urban areas, which can be masked with city or county-level data. The choropleth maps (Figures 6, S5-S11) highlight this variation and offer a visual representation of intra-urban heat islands, which studies have shown are remnants of prior redlining from the 1930s.^16,17^

Extreme heat events are becoming more frequent and intense with climate change.^1,2^ Not only do heat waves augment UHI intensity, but the UHI effect itself has been shown to cause additional hot days and heat waves in urban areas.^8,18^ This study further supports the idea that increasing UHI intensity due to climate change may increase heat stress and incidence of CVD in urban areas.^4^ The data also suggest that this burden is unequal – social factors such as a person’s race, ethnicity, and socioeconomic status may make them even more vulnerable to CVD.

There are several limitations to this study. Foremost, the CVD incidence data from CalEnviroScreen 4.0 was averaged over 2015-2017, whereas the UHII from CalEPA was averaged over 2006-2013. It is possible that values of UHII have changed between 2013 and 2017.

According to a global-scale UHII dataset spanning 2001-2020, there has been an interannual increase in UHII in most cities, and areas with higher UHII have a faster increase in UHII over time.^19^ Additionally, since California data was used in this study, it is unclear if these conclusions can be applied to other areas of the U.S. or more globally. This study also does not consider that individuals spend time outside of their home census tract, where they may be exposed to different UHIIs, or even the variation in UHII within census tracts. As well, the demographic data was derived from the ACS and is not directly linked to the CVD incidence data from CalEnviroScreen 4.0. This allowed for analysis of general trends in each census tract, however performing subgroup analyses for different racial and ethnic groups would not be reliable. This is an important future direction given the differences in CVD prevalence and risk factors amongst POC.^10^ It would be beneficial to research a study population in which the demographic information of the subjects is known, though this data is not publicly available on a census-tract level due to privacy concerns.

This study emphasizes the need for change in clinical practice and public health in the pursuit of environmental justice. It is important for clinicians to be aware of the impact of the UHI effect on health and risk for CVD. Practitioners should consider asking their patients about their living and work environments and if they have access to cooling measures. On a public health-level, this data can be used to identify census-tracts that may carry a higher heat stress burden.

Strategies to mitigate heat stress that have been shown to decrease mortality, such as distribution of home air-conditioners or access to cooling centers, can be implemented in vulnerable communities.^20^ However, these are not long-term solutions, since air-conditioning causes harm by generating heat outdoors, thereby increasing the UHI effect.^1^

More emphasis needs to be placed on preventative interventions that aim to decrease UHI intensity. These include implementing reflective roofing and pavement to decrease heat absorption and creating urban green spaces, green roofs, and vertical green walls with vegetation to augment shade and evaporative cooling.^21, 22^ While increased vegetation offers many benefits, newly planted trees may take decades to create adequate shade, and some urban areas are limited by water availability. It may be necessary to pair such long-term strategies with the installation of shade structures – preferentially with materials that minimize heat absorption – to reduce heat stress from mean radiant temperature.^23,24^

## Data Availability

All data is publicly available

https://oehha.ca.gov/calenviroscreen/report/calenviroscreen-40

https://calepa.ca.gov/climate/urban-heat-island-index-for-california/

## Acknowledgements

The authors would like to thank Dr. Nicholas Jackson for his statistical consultative contribution to this study.

